# A rare ORAI1 missense variant associates with risk of vascular diseases in White British adults

**DOI:** 10.1101/2024.03.20.24304648

**Authors:** Heba Shawer, Chew W Cheng, Karen E Hemmings, Abeer M Aldawsari, Gonzalo Revilla-González, Fabio Stocco, Jian Shi, David J Beech, Marc A Bailey

## Abstract

**Background:** Pathological remodelling of native vascular smooth muscle cells (VSMC) within the arterial wall is a key contributor to vascular disease. A driver of this remodelling is platelet-derived growth factor BB (PDGF-BB) and its signalling via activation of the calcium ion channel, ORAI1. Here, we investigated if there are associations of *ORAI1* polymorphisms with human cardiovascular disease.

**Methods and results:** We conducted candidate gene association analysis and revealed that a missense *ORAI1* variant (rs3741596, S218G) associates with an increased risk of hospital-diagnosed peripheral vascular disease, generalised atherosclerosis, acute ischaemic heart disease, and atrioventricular and left bundle-branch block in White British UK Biobank participants. Rs3741596 is also associated with higher circulating platelet counts and reduced total triglyceride levels. Functional analysis of the effects of rs3741596 S218G variant on ORAI1 channel function, via introduction of the S218G ORAI1 variant in HEK293 cells using CRISPR/Cas9 and investigation of its effects on SOCE, showed significantly enhanced SOCE compared to wild type cells, suggesting that the S218G variant enhances ORAI1 function.

**Conclusions:** Our results reveal an association between an *ORAI1* missense variant and occlusive vascular diseases. These findings provide a novel insight into the role of ORAI1 in vascular remodelling and highlight its potential as a treatment target for vascular pathologies.

## Introduction

Cardiovascular disease (CVD), encompassing occlusive atherosclerotic vascular diseases of the coronary, cerebral or peripheral arterial beds, vascular aneurysms, heart diseases (e.g., heart failure and conduction defects), hypertension, and stroke, remain the most lethal diseases worldwide, accounting for over a third of global deaths in 2019 ^1^. CVD is heavily driven by accumulation of atherogenic lipids in the arterial wall, thrombosis, and metabolic disease (e.g., obesity and diabetes). Genetic studies have advanced our understanding of the aetiology of CVD and helped identify effective therapeutic targets. For instance, discovering the associations of low-density lipoprotein receptor (LDLR) and Proprotein convertase subtilisin/kexin type 9 (PCSK9) polymorphisms with altered cholesterol homeostasis and increased risk of CVD led to the development of statins and PCSK9 inhibitors, that have proven to be effective in CVD risk reduction and the avoidance of major adverse cardiovascular events (MACE) ^2,3^. Candidate gene studies have revealed the association between mutations within the angiotensin converting enzyme (ACE) gene and increased risk of coronary heart disease ^4^, as well as the association between coagulation factor VII polymorphisms and myocardial infarction ^5^; findings that have led to the development of new effective therapies.

ORAI1 is a Ca^2+^ channel discovered in 2006 as the *de facto* pore-forming subunit of the Ca^2+^ release-activated Ca^2+^ (CRAC) channels, and has emerged as a promising therapeutic target, because of its accessibility to extracellular pharmacological inhibition as well as the success of ORAI1-targeted approaches in clinical trials. Targeting ORAI1 has already been clinically studied in inflammatory diseases and COVID-19-associated severe pneumonia ^6,7^. ORAI1 was originally linked with severe combined immunodeficiency in humans but there is now growing experimental evidence for a role in CVD ^8–12^. ORAI1 was also implicated in VSMC remodelling and vascular pathologies highlighting the therapeutic potential of targeting ORAI1 in occlusive vascular diseases^13^. ORAI1 genetic variants were previously found to be associated with susceptibility to Kawasaki disease ^14–16^, the most common cardiovascular disease in childhood, suggesting a potential link between ORAI1 genetic polymorphisms and cardiovascular pathology. Nonetheless, the potential association between ORAI1 polymorphisms and susceptibility to CVD in adults is still unknown.

Here, we studied ORAI1 as a promising druggable candidate protein ^17,18^ for CVD by investigating whether polymorphisms within the ORAI1 gene are associated with incidence of heart and vascular diseases in adults. We performed a candidate gene association analysis of the genetic alterations of *ORAI1* and their potential association with the risk of CVD and related metabolic diseases in UK Biobank participants from White British background. We revealed a novel association between a rare *ORAI1* missense variant and vascular diseases.

## Methods

### Participants

ORAI1 rare variants with minor allele frequency (MAF) less than 0.1% were examined for their association with cardiovascular and metabolic traits and disorders in UK Biobank participants. UK Biobank has ethical approval from the North-West Centre for Research Ethics Committee. This work accessed the UK Biobank anonymised data under application number 60315. The UK Biobank dataset comprises 487,409 participants aged 50-86 years, of which 430,628 are of White British ancestry. As most participants are of British descent, the analysis was performed only on individuals of British ancestry. The baseline characteristics of the study participants were obtained via a self-reported questionnaire. Participants’ characteristics are summarised in Table 1.

**Table 1:** Basic characteristics of the study participants.

| **Characteristic** | **Participants (n = 430,628)** |
| --- | --- |
| Mean age, years ± SD | 69.0 ± 8 |
| **Gender** |  |
| Female, n (%) | 233,062 (54.1%) |
| Male, n (%) | 197,566 (45.9%) |
| **Co-morbidities** |  |
| Stroke, n (%) | 6,100 (1.4%) |
| Hypertension, n (%) | 116,596 (27.1%) |
| Diabetes, n (%) | 18,073 (4.2%) |
| Asthma, n (%) | 51,015 (11.8%) |
| Heart/cardiac problem, n (%) | 1,554 (0.4%) |
| Angina, n (%) | 14,365 (3.3%) |
| Myocardial infarction, n (%) | 10,522 (2.4%) |

n, number of individuals; SD, standard deviation.

### Candidate gene association analysis

Imputation of genomic data was performed by UK Biobank ^19,20^, and we excluded samples with imputation quality les s than 0.4. Additional quality measures were applied to exclude participants with more than 10% missing data and variants with Hardy-Weinberg equilibrium (HWE) less than 1×10^-6^. We examined the association between 90 filtered rare variants within the ORAI1 gene and cardiovascular and metabolic disorders in UK Biobank participants, using logistic regression via PLINK version 2.0 software ^21^. Variants were annotated in hg19 coordinates using ANNOVAR ^22^. Cases were defined as individuals with primary diagnosis (UK Biobank Data-Field 41202) of CVD or metabolic disorders using the International Classification of Diseases (ICD-10) codes. To examine the association with blood lipids and haematological traits, linear regression model was used via PLINK version 2.0 software ^21^. Blood samples were collected and examined by the UK Biobank as described in ^23,24^. Covariates adjustment for the effects of sex, age, and the first ten principal components of population structure variation was performed. Regional association plots were generated using LocusZoom (http://locuszoom.sph.umich.edu) ^25^. This work was performed on ARC3, part of the High-Performance Computing (HPC) facilities at the University of Leeds. Data from the UK Biobank are available by application to the UK Biobank (https://www.ukbiobank.ac.uk/).

### CRISPR/Cas9-mediated ORAI1 point mutation

Human embryonic kidney 293 (HEK293) cell line with c.A658G point mutation in the ORAI1 gene was created using CRISPR/Cas9 mutagenesis and purchased from Ubigene Biosciences. Guide RNAs (sgRNA) with sequences (gRNA1: GTTGGCTGCTGCGCCACTGGCGG, gRNA2: GACGTTGGCTGCTGCGCCACTGG) were utilised as a template for homology-directed repair. The c.A>G (rs3741596, AGT>GGT) mutation was introduced by homology-directed repair, and a silent mutation (GGC>GGT) was introduced to avoid the binding and re-cutting of the sequence by gRNA after homology-directed repair.

### Cell Culture

HEK293 cells (ATCC, Teddington, UK) and HEK293 c.A658G mutant cells purchased from Ubigene Biosciences (Guangzhou, China) were cultured in Dulbecco’s Modified Eagle’s Medium (DMEM), supplemented with 10% v/v heat-inactivated foetal bovine serum (FBS) and 1% penicillin/streptomycin at 37°C in a humidified incubator at 5% CO_2_. Experiments were performed on cells up to passage 30. Experiments performed on cells at 95% confluence.

### Fura-2 Ca^2+^ addback assay

Fluorescence assays were performed using a FlexStation III (Molecular Devices) running Softmax Pro version 7.0 software. Cells were loaded with Fura-2AM (Molecular Probes, Thermo Scientific, UK) in 1.5 mM Ca^2+^ SBS containing 0.01% pluronic acid as an aid to Fura-2 uptake for 1 hour at 37 °C protected from light. Cells were washed 3 times with 1.5 mM Ca^2+^ SBS and then incubated in the presence of 1μM Thapsigargin in 0 mM Ca^2+^ SBS. Control wells were treated with an equivalent concentration of the vehicle DMSO. Cells were incubated for a further 30 minutes at room temperature protected from light. The Ca^2+^ addback was performed by the FlexStation III such that a final concentration of 0.3 mM Ca^2+^ was achieved. Fluorescence recordings were collected every 5 seconds for a total recording time of 230 seconds using 340/380 nm excitation and 510 nm emission wavelength. In experiments where JPIII was tested it was added during the store depletion phase of the experiment and remained at steady concentration during the Ca^2+^ add back phase. IC_50_ was determined by testing a range of JPIII concentrations and plotting a modified Hill equation.

### Statistical analysis

The associations of variants within ORAI1 (chr12:122,064,455– 122,080,583, GRCh37/hg19) with cardiovascular and metabolic disorders were examined using logistic regression, and the associations with blood lipids and haematological traits were examined using linear regression via PLINK version 2.0, after adjusting for effects of sex, age, and the first ten principal components that conveys variations in population structure. Bonferroni correction was applied on the P-value to adjust for the number of variants tested. Nominal P-value of 0.05 was used to indicate statistically significant associations for the candidate gene association analysis. The ratio between excitation at 340 nm and 380 nm was calculated (ΔF^340/380^). The first six readings were used to calculate the baseline which was subtracted to determine the baseline corrected ΔF^340/380^. The peak baseline corrected ΔF^340/380^ for each treatment was analysed by One way ANOVA with Tukey’s post hoc test. A p-value of <0.05 was considered significant.

## Results

### ORAI1 and atherosclerotic cardiovascular diseases

To study the potential association between *ORAI1* variants and risk of CVD, we first tested the association between ORAI1 variants and atherosclerotic CVD in UK Biobank White British participants. We then took forward the non-synonymous ORAI1 variant that reached nominal significance of P<0.05 for investigation of association with risk of additional cardiovascular and metabolic traits and association with alterations in blood lipid and haematological parameters in this population. We focused on studying the associations of rare ORAI1 variants. Unlike the common variants, rare variants are more likely to be associated with a large difference in disease risk and more likely to be associated with altered protein function. The association between 90 rare ORAI1 variants and the incidence of cardiovascular diseases in 430,628 UK Biobank participants from White British ancestry aged between 50-86 years was investigated. Summary of the participants’ characteristics and medical history (self-reported by the participants in an interview by a trained nurse) is presented in Table 1.

We identified 17 rare variants with minor allele frequency (MAF) less than 0.1% associated with hospital diagnosed peripheral vascular disease (PVD). Furthermore, 35 and 20 ORAI1 variants were found to be associated with generalised atherosclerosis and acute ischaemic heart disease, respectively. These variants include only one non-synonymous SNP (rs3741596), which is located at exon 2 of the ORAI1 gene (NM_032790.3:c.A652G, Figure 1A), and was further analysed for association with other traits associated with CVD and metabolic disorders. The ORAI1 rs3741596 nonsynonymous SNP is associated with hospitalisation for PVD, with a nominally increased risk in the variant carriers (n = 860, OR= 1.7 with 95% CI = 1.1-2.7, P = 0.012). Similar to this increased risk of hospitalisation for PVD, the hospitalisation or death due to PVD (n = 1156, OR = 1.7 with 95% CI = 1.2-2.5 P = 0.004) was also more common in the carriers of the rs3741596 variant. Carriers of the minor allele were also shown to have 6.8-fold higher risk (OR 95% CI = 2.2-21.2, P= 0.001) of hospitalisation for generalised atherosclerosis (n=33), relative to the control group. Similarly, rs3741596 was associated with increased risk of acute ischaemic heart disease (n = 436, OR = 1.8 with 95% CI = 1-3.3, P = 0.0497, Supplementary Table 1). The lists of the nominally significant SNPs associated with PVD, generalised atherosclerosis and acute ischaemic heart disease are shown in Supplementary Tables 2-4. The distribution of the minor allele of this rs3741596 variant among the different ethnic populations, from the 1000 Genomes Project phase 3, showed high MAF in the East Asian compared to the other populations. In East Asian population of the 1000 Genomes Project, the frequency of the G allele of the rs3741596 variant was 11.3%; while its frequency in the African, American, European, and South Asian populations were 7.9%, 0.4%, 0.9% and 0.7%, respectively ^26^. These data suggest that the minor allele (G) of the rs3741596 variant is rare in the American, European, and South Asian, while more frequent in the East Asian and African populations.

**Figure 1:**
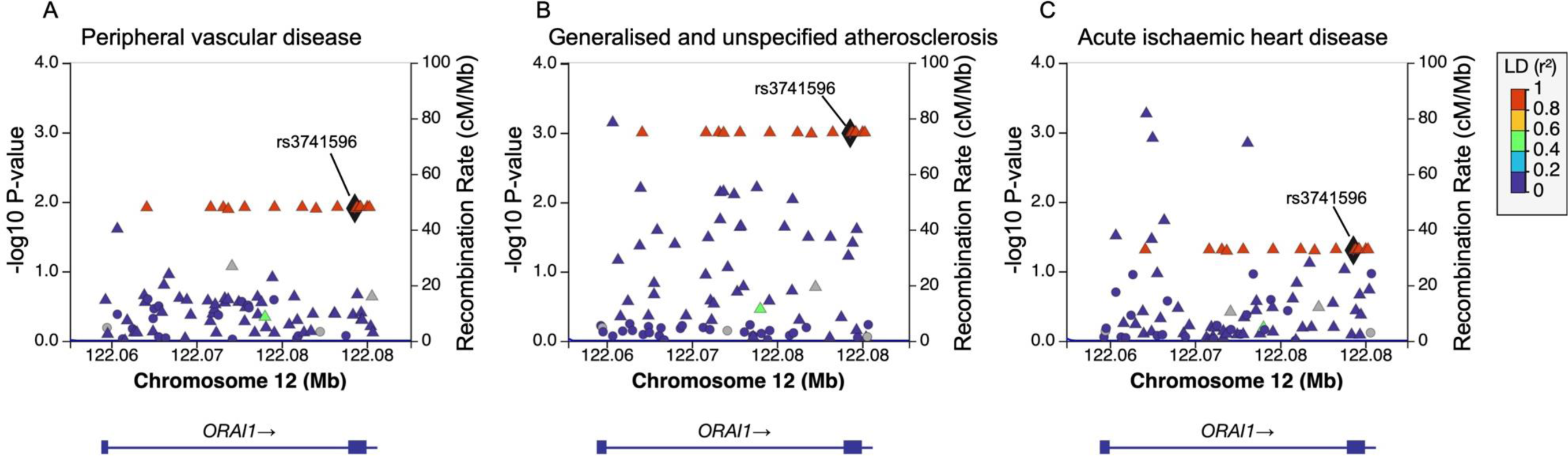
Regional association plots presenting SNPs within the ORAI1 locus and their associations with peripheral vascular disease, generalised and unspecified atherosclerosis, and acute ischaemic heart disease. Variants are plotted according to the −log10 scale of the P-value of their association with peripheral vascular disease **(A)**, generalised and unspecified atherosclerosis **(B)**, and acute ischaemic heart disease **(C)** displayed on the left y-axis. The right y-axis shows the recombination rate estimated from the 1000 Genomes European data, and the x-axis displays the chromosomal locations of the variants on build GRCh37/hg19. Variants are color-coded based on their level of linkage disequilibrium (LD, r2 values) with rs3741596.

### ORAI1 and cardiac conduction disorders

We and others have previously reported the implications of SOCE and ORAI1 in cardiac pathologies, including progressive left ventricular systolic dysfunction, cardiac hypertrophy, and arrhythmia ^11,27–30^. We therefore investigated the potential association between the ORAI1 missense variant rs3741596 and cardiac conduction disorders. Our candidate gene association analysis revealed novel association between the ORAI1 nonsynonymous variant, rs3741596, with 1.9 odds ratio (95% CI = 1.4-2.7, P = 0.0002) of hospitalisation for atrioventricular and left bundle-branch block (n = 1233, Supplementary Tables 5 and 6). rs3741596 showed no significant association with atrial fibrillation and flutter (n = 9657, P = 0.85), paroxysmal tachycardia (n = 2461, P = 0.19), or cardiac arrest (n = 304, P = 0.54).

### Obesity, diabetes, and blood lipids traits

As obesity, diabetes and dyslipidaemia are risk factors for CVD, we investigated the association between rs3741596 and these metabolic traits that could provide an insight into the aetiology of the association between ORAI1 polymorphisms and CVD. rs3741596 did not show any association with obesity (n = 617, P = 0.27) or insulin dependent diabetes mellitus (n = 747, P = 0.1) (Supplementary Table 7). We also tested rs3741596 for association with circulating lipid phenotypes in White British UK Biobank participants (n = 103,731) and observed nominal association with lower total triglycerides (β = −0.035, P = 0.017), and lower triglyceride content in VLDL (β = −0.03, P = 0.02), LDL (β = −0.002, P = 0.02), and HDL (β = −0.003, P = 0.03). There was no association observed with either circulating total cholesterol, non-HDL cholesterol, remnant cholesterol (non-HDL and non-LDL cholesterol), VLDL cholesterol, LDL cholesterol, or HDL cholesterol. We observed associations between rs3741596 and reduced concentration of chylomicrons and extremely large VLDL particles (β = −0.0000001, P = 0.0075), as well as of its structural components, including, the total lipids (β = −0.013, P = 0.01), phospholipids (β = −0.002, P = 0.006), cholesterol (β = −0.003, P = 0.01), and cholesteryl esters (β = −0.001, P = 0.02) carried by the chylomicrons and extremely large VLDL. Nominal association of rs3741596 with reduction in phospholipids (β = −0.009, P = 0.05) and total lipids (β = −0.05, P = 0.03) in the VLDL fraction was also observed (Figure 2, Supplementary Table 8). These findings highlight a potential involvement of ORAI1 in triglycerides and lipid homeostasis beyond the parameters targeted by current best medical therapy.

**Figure 2:**
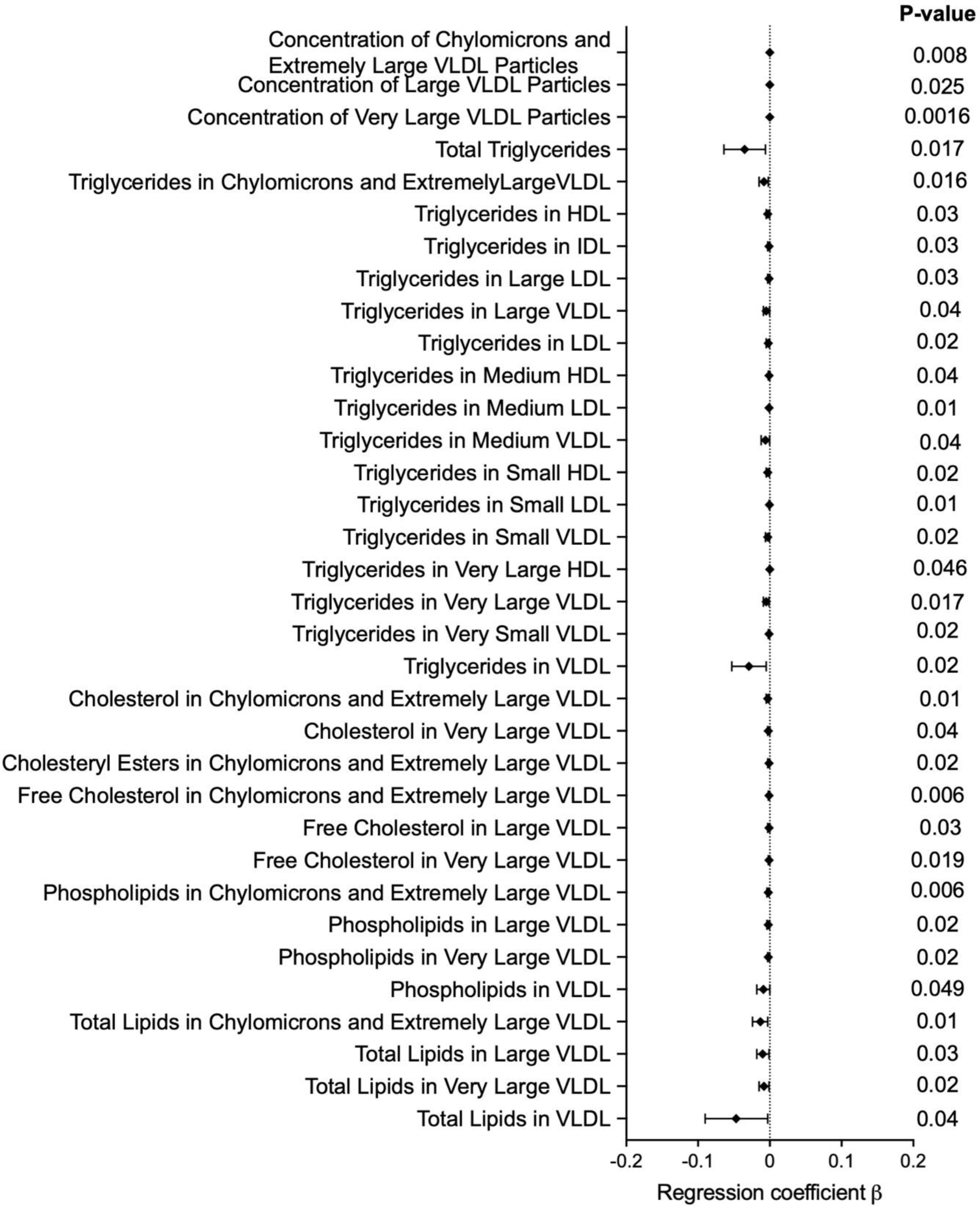
Forest plot demonstrating significant associations of rs3741596 with blood lipids traits. The diamonds and horizontal lines represent the regression coefficient (β) and 95% CI, respectively.

### ORAI1 and blood cell traits

As blood cells play a crucial role in vascular homeostasis and athero-thrombotic vascular diseases, we investigated the potential association between the ORAI1 missense mutation, rs3741596, and 30 blood cell phenotypes (Figure 3, Supplementary Table 9). For platelet indices, we observed nominally significant associations between rs3741596 and elevated platelet count (n = 417,730, β = 1.8, P = 0.016), and with reduced mean platelet (thrombocyte) volume (n = 417,725, β = −0.038, P = 0.008) and platelet distribution width (n = 417,725, β = −0.016, P = 0.015). Previous studies have shown that ORAI1 is expressed in human platelets and ORAI1 variants were previously identified to be associated with defects in platelet activation and thrombocytopenia ^31,32^. These findings highlight the role of ORAI1 in platelet function, which dysregulation contributes to the pathogenesis of vascular diseases, including atherosclerosis and Neo-Intimal Hyperplasia (NIH). For red blood cell traits, the missense variant rs3741596 was nominally associated with red blood cell (erythrocyte) distribution width (n = 417731, β = 0.03, P = 0.007). Finally, nominal association was also observed with reduction in lymphocyte percentage in leukocytes (n = 417000, β = −0.2, P = 0.02).

**Figure 3:**
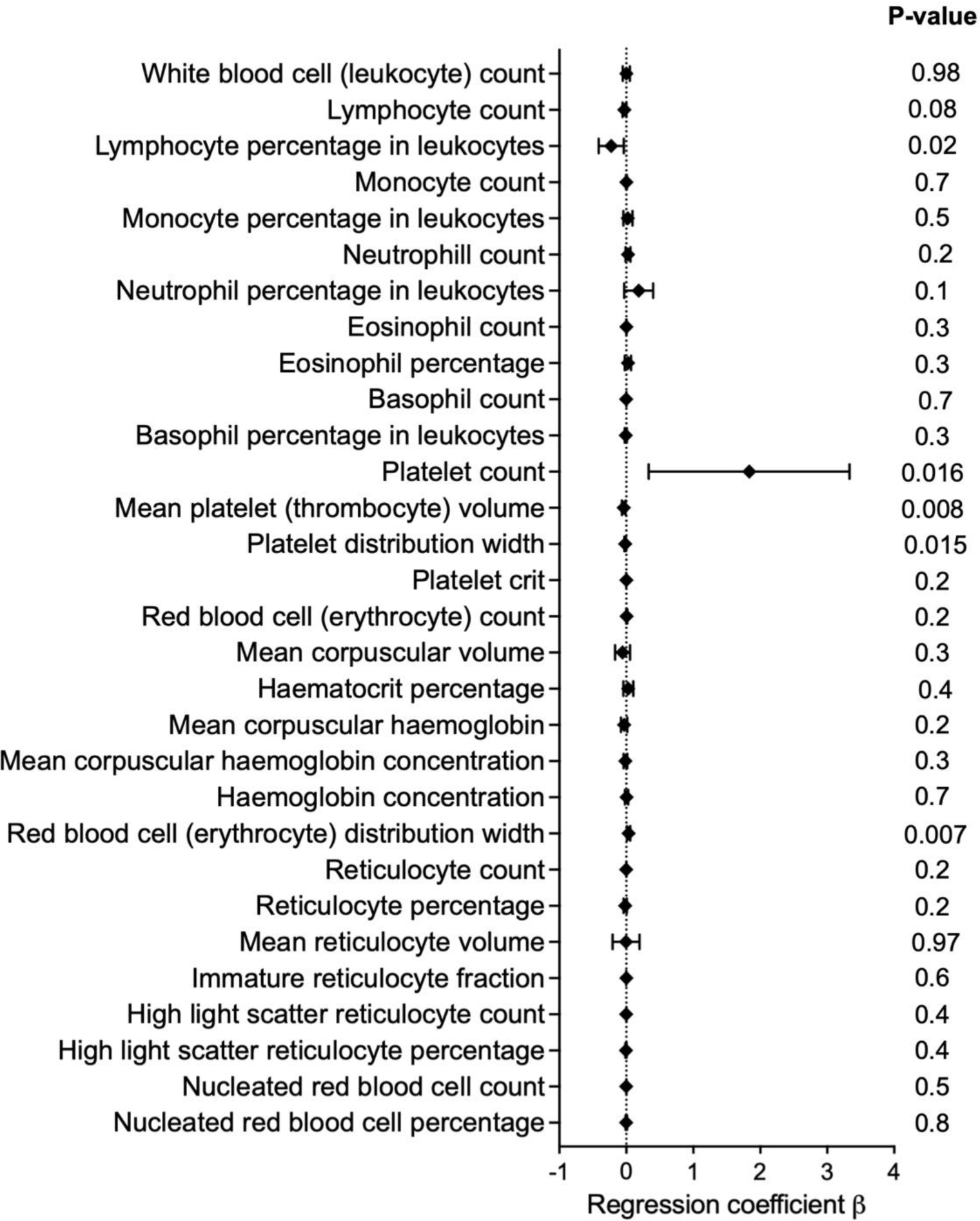
Forest plot demonstrating the associations of rs3741596 with blood cell traits. The diamonds and horizontal lines represent the regression coefficient (β) and 95% CI, respectively.

### Functional validation of the candidate ORAI1 missense variant

To study the effect of the rs3741596 S218G variant on ORAI1 channel function, we introduced the A652G ORAI1 variant in HEK293 cells using CRISPR/Cas9 and investigated its effects on SOCE. For this, we induced internal Ca^2+^ store depletion by treating cells with the Sarco/endoplasmic reticulum Ca^2+^-ATPase (SERCA) inhibitor, thapsigargin (TG), under Ca^2+^-free conditions. This was followed by extracellular Ca^2+^ addback that activates SOCE. SOCE was then observed as an elevation of cytosolic Ca^2+^ levels following the extracellular Ca^2+^ addback. SOCE in HEK293 cells carrying the S218G ORAI1 mutation was assessed using fura-2-based Ca^2+^ imaging and was shown to be significantly enhanced compared to wildtype cells, suggesting that the S218G variant enhances ORAI1 function (Figure 4). We also tested our potent and specific ORAI1 small-molecule inhibitor, JPIII against the S218G ORAI1 mutant and found it to have similar potency (ORAI1 S218G mutant IC_50_ = 33 nM vs.29 nM for WT HEK cells in the same assay).

**Figure 4:**
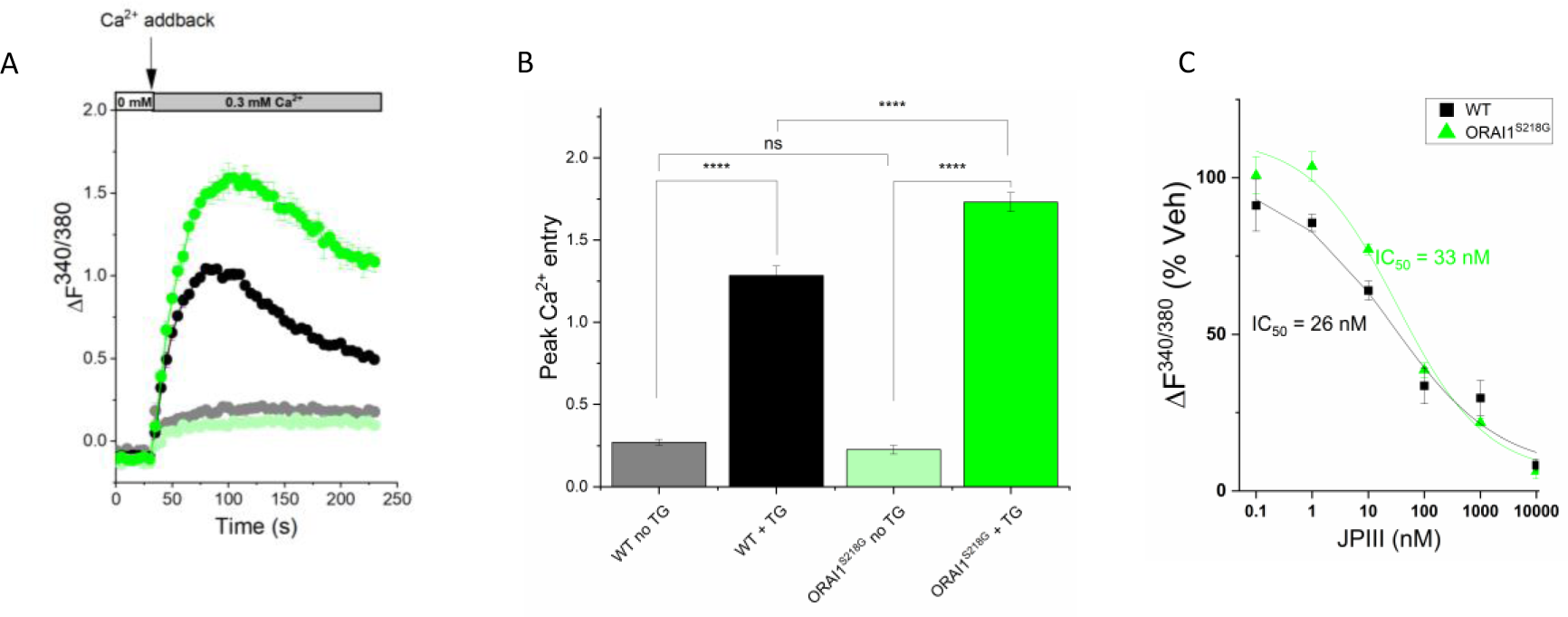
Functional characterisation of the ORAI1 S218G mutation in the Ca^2+^ addback assay. **A,B)** Representative fluorescence over time graph of thapsigargin (TG) induced store operated Ca^2+^ entry. Data are baseline corrected. TG is added before recording. Ca^2+^ addback time = 30 seconds (s). Data presented as mean ± SEM (n=4). **B)** Quantification of peak Ca^2+^ entry in wild type HEK293 cells and cells with the S218G variant. Data presented as mean ± SEM (N=16). **C)** IC_50_ curve for inhibition of ORAI1 by JPIII in wild type HEK293 cells and cells with the S218G variant (n=3). ORAI1 S218G HEK293 cells (green), WT HEK293 cells (black) and control cells not exposed to TG; ORAI1 S218G HEK293 cells (light green), WT HEK293 cells (grey). Statistical significance using one-way ANOVA with Tukey’s post hoc test ns = not significant, **** P<0.0001

## Discussion

Despite the increased utilisation of robust primary and secondary prevention strategies and the reduction in major adverse cardiovascular events, CVD remains a significant clinical problem and new therapeutic approaches are needed. The identification of a “druggable” target that modulates the phenotypic remodelling of VSMC to reduce atherosclerotic burden and reduce the risk of restenosis following vascular interventions is of high priority. We identified an association between a rare gain of function ORAI1 missense variant and risk of hospitalisation for PVD, generalised atherosclerosis, acute ischaemic heart disease and atrioventricular and left bundle-branch block in White British UK Biobank participants.

Genetic loss of function mutations within ORAI1 that result in ORAI1 channelopathy have previously been reported in patients with immunodeficiency, ectodermal dysplasia anhidrosis (EDA), and muscular hypotonia ^31,33,34^. Whereas the main manifestation of ORAI1 gain-of-function mutations was tubular aggregate myopathy (TAM) and Stormorken syndrome ^35,36^. ORAI1 genetic variants were also found to be associated with susceptibility to Kawasaki disease, which is the leading cause of CVD in children ^14–16^. In animal models, we have previously identified a cardio-protective effect of pharmacological or transgenic ORAI1 inhibition after pressure overload in mice ^11^, favourable effects of ORAI1 *in vivo* inhibition in pulmonary arterial hypertension ^12^, and others have reported beneficial effects of ORAI1 inhibition or gene silencing in atherosclerosis ^10^ and NIH ^8,9^. Although these studies support involvement of ORAI1 signalling in cardiovascular pathologies in model systems the significance of these observations to human patients with CVD were unclear.

This study reveals a novel association of a rare missense variant, rs3741596 (c.A652G), within the ORAI1 gene with generalised atherosclerosis, acute ischaemic heart disease, hospitalisation for PVD, and atrioventricular and left bundle-branch block in White British UK Biobank participants. Additionally, we found associations of rs3741596 with elevated circulating platelet counts (β = 1.8, P = 0.016), reduced mean platelet (thrombocyte) volume (β = −0.038, P = 0.008), lower total triglyceride levels (β = −0.035, P = 0.017) but without association with obesity or diabetes, which are important risk factors for atherosclerotic vascular diseases. ORAI1 expression was previously reported in hematopoietic cells, including platelets ^32^, and a role of ORAI1-mediated SOCE in platelet activation was identified, whereby defective SOCE in Orai1^-/-^ or Stim1^-/-^ mice resulted in impaired platelet activation and thrombus formation ^37–39^. It was also previously reported that patients heterozygous for the loss of function R91W or G98S ORAI1 mutation were presented with low platelet counts ^40^. Furthermore, the introduction of single nucleotide polymorphism in STIM1, which impaired its activation in response to ER Ca^2+^ deletion, resulted in macrothrombocytopaenia, impaired platelet activation and bleeding disorder in mice ^41^, and gain-of-function mutations in STIM1 were observed in patients with York Platelet syndrome ^42^ and patients suffering from thrombocytopenia ^43^. These findings highlight the role of ORAI1 in platelet function and suggest a potential involvement in platelet activation and adhesion in atherosclerosis. Our findings of rs3741596 association with elevated circulating platelet counts suggest that ORAI1 activity does not only influence platelet function but is also associated with altered platelet counts. Interestingly, rs3741596 was among the ORAI1 variants previously reported to be associated with Kawasaki disease ^14–16^, supporting its implication in cardiovascular pathologies. It was also reported to be associated with susceptibility to atopic dermatitis, an inflammatory skin disease ^44^. The association of rs3741596 with inflammatory skin diseases ^44^ and Kawasaki disease ^14–16^, supports the role of this variant in inflammatory and cardiovascular diseases.

The rs3741596 variant is located within exon 2 of ORAI1 (NM_032790.3:c.A652G), altering the reference nucleotide [A] at position 652 of the ORAI1 transcript to the nucleotide [G], leading to an alteration of [AGT] codon to [GGT]. This variant, therefore, leads to a Serine (S) to Glycine (G) substitution at position 218 of the ORAI1 protein (NP_116179.2: p.S218G). This S218G mutation is located within the second extracellular loop of the ORAI1 channel, a region with unclear function. The region of the second extracellular loop around the S218G mutation was shown to have low conservation across species ^15^ and is predicted by Clinvar (allele ID: 372869) to be likely benign ^45^. It was previously proposed that this S218G mutation is potentially a gain-of-function mutation ^14^. Our finding of increased SOCE in cells with the rs3741596 variant (S218G) compared to wildtype cells supports this observation (Figure 4). Another study that analysed the effects of the rs3741596 variant suggest that the S218G ORAI1 mutation results in increased plasma membrane ORAI1 abundance and a subsequent mismatch in the coupling ratio of ORAI1 and STIM1 ^46^.

The identification of genetic polymorphisms that associate with an increased risk of CVD could help unravel novel therapeutic targets and improve our understanding of the disease and provide the starting points for development of additive therapeutics for patients with CVD. Our findings highlight the potential implication of ORAI1 activity in the pathogenesis of occlusive vascular diseases. Our analysis focused on the non-synonymous ORAI1 variant found to be associated with vascular diseases. Therefore, further analysis of the silent and intronic SNPs associated with vascular diseases is still needed. Furthermore, the study was limited to participants from White British (Caucasian) background, and therefore our findings do not represent the wider population. Further studies should investigate the genetic association of ORAI1 variants and CVD in a larger and more diverse population.

Taken together, we demonstrated the novel association of the missense ORAI1 rs3741596 variant with increased risk of hospitalisation for PVD, generalised atherosclerosis, acute ischaemic heart disease and atrioventricular and left bundle-branch block, as well as association with blood lipids and haematological traits. We were able to demonstrate that the rs3741596 results in enhanced SOCE. The results presented in this study provide insight into the role of ORAI1 in vascular remodelling and the possible therapeutic potential of pharmacologically targeting ORAI1 for pathologic vascular remodelling.

## Data Availability

All data is provided in the supplementary materials online.

## Acknowledgments

This work was conducted using UK Biobank datasets. We thank the UK Biobank participants and staff. This work was undertaken on ARC3, part of the High-Performance Computing (HPC) facilities at the University of Leeds, UK.

## Supplementary files

Supplementary Table 1: Description of the cardiovascular disease outcomes and their associations with the ORAI1 nonsynonymous SNP, rs3741596.

Supplementary Table 2: Description of cardiac conduction traits and their association with the ORAI1 nonsynonymous SNP, rs3741596.

Supplementary Table 3: The association of rs3741596, with obesity and insulin dependent diabetes mellitus.

Supplementary Table 4: ORAI1 variants with MAF less than 0.1% associated with peripheral vascular disease.

Supplementary Table 5: ORAI1 variants with MAF less than 0.1% associated with generalised atherosclerosis.

Supplementary Table 6: ORAI1 variants with MAF less than 0.1% associated with acute ischaemic heart disease.

Supplementary Table 7: ORAI1 variants with MAF less than 0.1% associated with atrioventricular and left bundle branch block.

Supplementary Table 8: Association results of rs3741596 with circulating lipids traits in UK Biobank.

Supplementary Table 9: The associations between the ORAI1 nonsynonymous SNP, rs3741596, and blood cell traits.

